# Evidence for a Causal Pathway Between Socioeconomic Status and Melanoma in Situ

**DOI:** 10.1101/2025.04.23.25326166

**Authors:** Umber Dube, Jennifer Y. Lin

## Abstract

**Importance:** The genetic architecture of disease risk may not be independent of social determinants. This can be leveraged to investigate for causality.

**Objective:** To investigate for a possible causal relationship between socioeconomic status (SES) and melanoma in situ (MIS) based on their genetic architectures.

**Design:** Genetic correlation study

**Setting:** Multicenter and population-based data sources.

**Participants:** The Ingold et al., 2024 GWAS summary statistic dataset is derived from 3,564 MIS cases, 10,552 invasive melanoma (MM) cases, and 1,022,070 melanoma-free controls. Individuals with both MIS and MM were only included as MM cases. The Kweon et al., GWAS summary statistic dataset on income, a proxy for SES, is derived from an effective sample size of 668,288 individuals.

**Main Outcome(s) and Measure(s):** Genetic correlation between MIS, MM, and SES.

**Results:** We obtained European genomic ancestries-based GWAS summary statistics for MIS (3,564 cases and 1,022,070 melanoma-free controls), MM (10,552 cases and 1,022,070 melanoma-free controls), and SES (668,288 individuals). We identify a positive and significant genetic correlation between MIS and SES (0.14, 95% CI 0.06 to 0.21; p = 5.55 × 10^-04^) but not MM and SES (0.05, 95% CI −0.01 to 0.11; p = 0.11). The genetic architecture of MIS subtracting that of MM (MIS-MM) remained positively and significantly correlated with the genetic architecture of SES (0.23, 95% CI 0.07 to 0.39; p = 4.18 × 10^-03^). In contrast, the genetic architecture of MM subtracting that of MIS (MM-MIS) is negatively correlated with the genetic architecture of SES (−0.24, 95% CI −0.42 to −0.06; p = 8.01 × 10^-03^). Finally, taking a Mendelian Randomization approach, we identify consistent evidence for a causal pathway between MIS and SES but not MM and SES.

**Conclusions and Relevance:** The genetic architecture of SES correlates with that of MIS but not MM. There is also evidence for a causal link between SES and MIS. Both these findings support an overdiagnosis of MIS. Importantly, our results demonstrate genetic risk scores for disease are not inherently independent of social determinants of diagnosis. Clinical application of genetics-based risk stratification without consideration of social determinants may have limited utility.

**Key Points:** *Question:* Is socioeconomic status genetically correlated with melanoma in situ and is there any evidence for a causal relationship?

*Findings:* In this genetic correlation study based on data from 3,564 melanoma in situ cases, 10,552 invasive melanoma cases, 1,022,070 melanoma-free controls, and 668,288 individuals with income data; we identify a positive and significant genetic correlation between socioeconomic status and melanoma in situ but not invasive melanoma. We also identify Mendelian Randomization-based evidence for a causal relationship between socioeconomic status and melanoma in situ but not invasive melanoma

*Meaning:* The genetic architecture of disease risk is not independent of social determinants of diagnosis. Clinical application of genetics-based risk stratification without consideration of social determinants may have limited utility.

## Introduction

Invasive melanoma (MM)-specific survival increases with earlier stage at detection.^1^ While efforts to identify disease earlier have been successful in increasing the number of melanoma in situ (MIS) diagnoses^2,3^, they have not improved melanoma-specific mortality.^4^ MIS can progress to invasive disease, but MIS is not an obligate precursor of MM.^5,6^ As such, MIS is at risk of overdiagnosis, which is the increased detection of low-risk disease. This occurs more commonly in those with increased access and utilization of healthcare, both of which correlate with socioeconomic status (SES).^7^ Indeed, individuals with a diagnosis of MIS typically have higher SES and demonstrate increased survival relative to both those with thin MM and the general population.^8^ Thus, MIS is likely overdiagnosed.

Genetics-based risk stratification has been proposed as a strategy to identify those at increased risk of melanoma mortality without exacerbating overdiagnosis.^9^ However, the utility of this approach is dependent on the genetic architecture of disease risk being independent from non-genetic factors influencing disease diagnosis. This can be tested empirically. Income – a proxy for SES – has genome wide association study (GWAS)-significant genetic loci.^10^ These can be leveraged to investigate genetic overlap and provide evidence for causality. Here we investigate the genetic relationship between SES, MIS, and MM.

## Methods

### Datasets analyzed and their standardization

We obtained European genomic ancestries-based GWAS summary statistics for MIS and MM from Ingold et al., 2024.^9^ Their study included 3,564 MIS cases, 10,552 MM cases, and 1,022,070 melanoma-free controls. Importantly, individuals with both MIS and MM were only included as MM cases. Similarly, we obtained income GWAS summary statistics from Kweon et al., 2025, which are derived from an effective sample size of 668,288 individuals.^10^ Given the strong correlation between the genetic architectures of income and education (0.92, 95% CI 0.91 to 0.93)^10^, we summarize this architecture as representing that of SES in the manuscript. Prior to genetic correlation analyses we standardized the datasets by annotating and processing all variants as previously described.^11^ In short, we used ANNOVAR^12^ software to assign each variant a rs-identifier based on dbSNP^13^ 151 and its effect allele a frequency based on the non-Finnish European population from gnomAD^14^ (v2.1.1). For instances where multiple variants shared the same rs-identifier we selected the variant supported by the largest effective sample size.

### Genetic correlation analyses

We used GNOVA software^15^ for our primary genetic correlation analyses. Prior to GNOVA analysis, we processed the standardized GWAS summary statistic datasets using the munge_sumstats.py tool from LDSC software^16^ with standard parameters. This removed variants with an annotated effect allele frequency less than 0.05, variants supported by a low sample size, strand ambiguous variants, and variants absent from the HapMap3^17^ study. We ran GNOVA software on these processed GWAS summary statistic datasets using standard parameters and the 1000 Genomes^18^ European population-based linkage disequilibrium reference data provided with the software. We present genetic correlation estimates corrected for sample overlaps by GNOVA. As a sensitivity analysis, we repeated the genetic correlation analyses with HDL^19^ software using the HapMap2 reference panel provided with the software. This reference panel was selected to maximize the overlap between standardized GWAS summary statistic variants and reference variants.

### GWAS-by-subtraction

We used GSUB software^20^ to calculate the signed differences between MIS and MM GWAS summary statistics. We ran GSUB using standard parameters assuming an equal prevalence of 0.0045 for both MIS and MM. Given the strong genetic correlation between MIS and MM, the effective sample sizes for variants of each of the resulting genetic architectures were small. Variants for which the effect allele frequency fell outside the range for stable calculation of effective sample size were assigned the median effective sample size from those that were calculable. We compared the genetic correlation between the calculated signed differences’ genetic architectures and SES using GNOVA software as above.

### Mendelian randomization and identification of pleiotropy

We performed Mendelian randomization (MR) analyses using the R packages MendelianRandomization^21^, TwoSampleMR^22^, and mr.raps^23^. Per published MR guidelines^24^, we selected the following MR methods for our analyses: inverse variance weighted with random effects (IVW), robust adjusted profile score (RAPS), weighted median, and constrained maximum likelihood-based (cML)^25^ and ran them using the default parameters. We selected SES-associated genetic variants as instruments for these analyses using PLINK 1.9^26^ software. Selection parameters included strength of association with SES (p-value < 5 × 10^-08^) and independence (r^2^ < 0.001) from variants in a surrounding 10,000 kilobase window based on 1000 Genomes European population. Pleiotropic variant associations were identified by querying the GWAS catalogue via LDTrait.^27^

### Statistical analyses

P values less than 0.05 are considered nominally significant through 2-sided significance testing and statistically significant if they are less than 0.05 divided by the number of independent tests conducted during each analysis.

## Results

*Positive genetic correlation between socioeconomic status and melanoma in situ but not invasive melanoma*.

We first confirmed the reported genetic correlation between MIS and MM (0.83, 95% CI 0.72 to 0.94; p = 3.85 × 10^-50^, Figure 1) using GNOVA software. We next investigated genetic correlation with SES. We identified a positive and significant correlation between the genetic architectures of SES and MIS (0.14, 95% CI 0.06 to 0.21; p = 5.55 × 10^-04^, Figure 1) but not that of SES and MM (0.05, 95% CI −0.01 to 0.11; p = 0.11, Figure 1). To validate our approach, we applied an orthogonal genetic correlation method: HDL. Out HDL genetic correlation results were consistent with those from GNOVA, identifying a positive and significant correlation between SES and MIS (0.17, 95% CI 0.05 to 0.29; p = 5.53 × 10^-03^) but not between SES and MM (0.16, 95% CI –0.36 to 0.68; p = 0.56).

**Figure 1.**
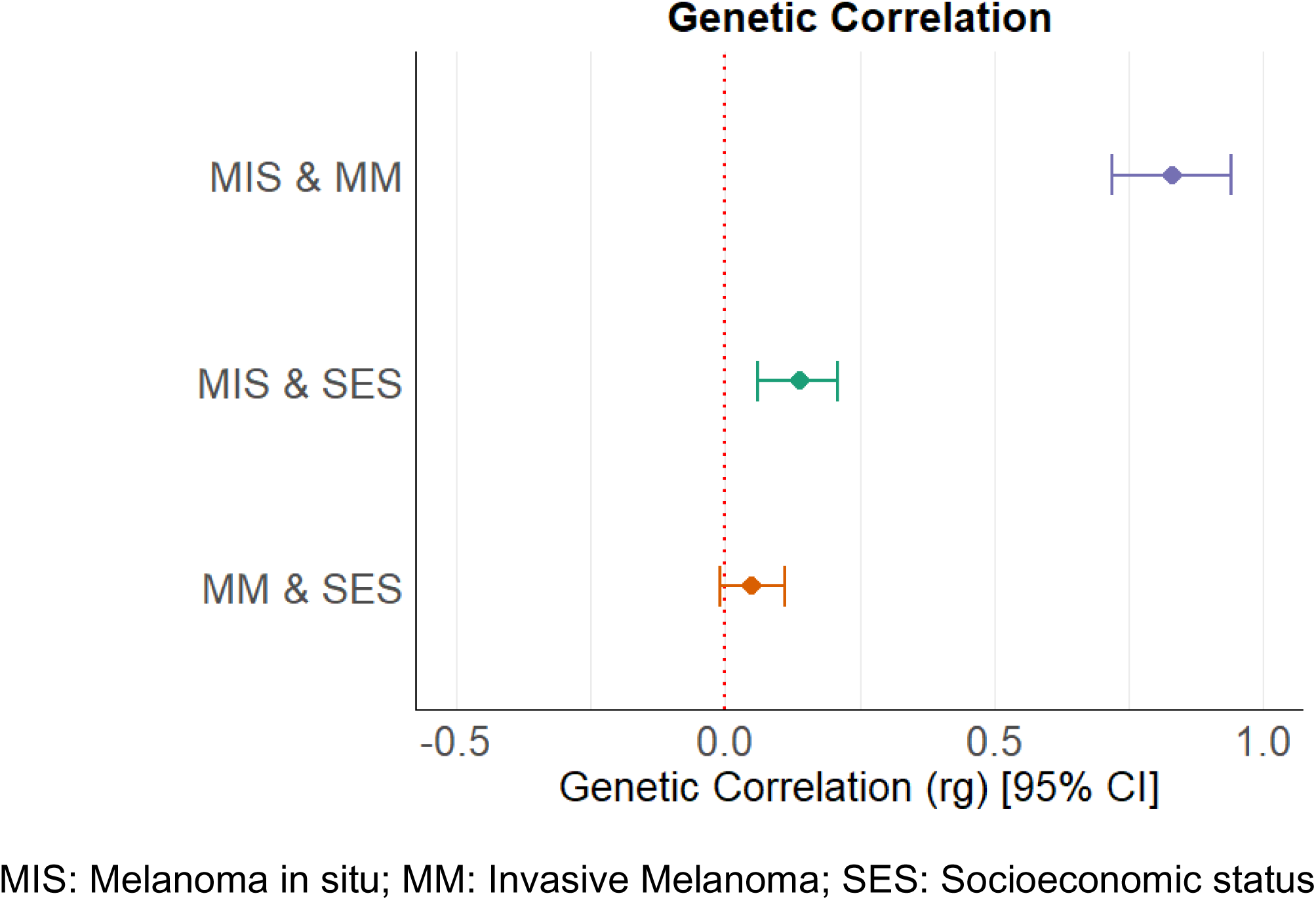
Genetic correlation between Socioeconomic Status, Invasive Melanoma, and In Situ Melanoma

### Comparative subtraction demonstrates consistent correlation between the genetic architecture of socioeconomic status and that of melanoma in situ independent of invasive melanoma

Given that MIS and MM demonstrate a differential genetic correlation with SES, despite their strong correlation with each other, we hypothesized that the genetic architecture of MIS independent of that from MM would demonstrate a stronger genetic correlation with SES than MIS alone. To investigate this, we calculated the signed differences between the genetic architectures of MIS and MM via GWAS-by-subtraction using GSUB software. We then performed genetic correlation analyses with SES. The residual genetic architecture of MIS after subtracting that of MM, which we term MIS-MM, maintained a significant and positive correlation with SES (0.23, 95% CI 0.07 to 0.39; p = 4.18 × 10^-03^). Notably, while the point estimate for genetic correlation with SES is greater for MIS-MM than for MIS alone, the statistical power loss from GWAS-by-subtraction results in overlapping confidence intervals. Conversely, MM-MIS – the residual genetic architecture of MM after subtracting that of MIS – was significantly negatively correlated with SES (−0.24, 95% CI −0.42 to −0.06; p = 8.01 × 10^-03^) and did not overlap with the point estimate for MM alone.

### Evidence for causality between socioeconomic status and melanoma is situ via Mendelian Randomization analyses

As our genetic correlation results suggested a causal pathway between SES and MIS, we took a Mendelian Randomization (MR) approach to investigate further. In brief, MR analyses provide evidence for a causal pathway based on taking genetic variants associated with an exposure as instrumental variables and testing if they are also associated with an outcome.^28^ Following guidelines for MR analyses^24^, we find consistent evidence for causality between MIS and SES (Table 1). This result was consistent through leave-on-out and single variant sensitivity analyses, bolstering confidence in this result (Supplementary Figures 1-2). In contrast, none of the MR methods applied to MM and SES yielded results that passed multiple test correction (Table 1). Sensitivity analyses demonstrated considerable heterogeneity, that was driven by a single variant: rs77270200 (Supplementary Figures 3-4), which is associated with increased risk for MM (beta = 0.39; 95% CI 0.34 to 0.44; p = 2.22 × 10^-54^)^9^ and MIS (beta = 0.39; 95% CI 0.08 to 0.26; p = 155 × 10^-04^)^9^ but decreased SES (beta = −0.02; 95% CI −0.03 to −0.01; p = 6.13 × 10^-12^)^10^. While this variant is pleiotropic and has been associated with multiple traits (Supplementary Table 1), its GWAS-significant association with MM limits its use as an MR instrumental variable. Excluding this variant from the MM and SES MR analysis provided more nominally significant results, but again none that passed multiple correction (Supplementary Table 2). Overall, we find consistent evidence to support causality between MIS and SES but not MM and SES.

**Table 1.** Mendelian Randomization investigating causality between SES and MIS versus MM.

| Mendelian Randomization Method | MIS | MM |
| --- | --- | --- |
| IVW | 0.52 [0.14, 0.90]<sup>a</sup><br>($6.72 \times 10^{-03}$) | 0.11 [-0.31, 0.53]<sup>b</sup><br>(0.60) |
| MR-RAPS | 0.56 [0.18, 0.94]<br>($4.14 \times 10^{-03}$) | 0.29 [-0.01, 0.60]<br>($6.18 \times 10^{-02}$) |
| Weighted Median | 0.88 [0.34, 1.42]<br>($1.37 \times 10^{-03}$) | 0.24 [-0.12, 0.61]<br>(0.19) |
| MR-cML | 0.58 [0.14, 1.02]<br>($9.02 \times 10^{-03}$) | 0.33 [0.04, 0.63]<br>($2.63 \times 10^{-02}$) |

<sup>a</sup>MIS IVW: Heterogeneity test statistic (Cochran's Q) = 146.2113 on 136 degrees of freedom, (p-value = 0.2596). $I^2$ = 7.0%. F statistic = 44.0.
<sup>b</sup>MM IVW: Heterogeneity test statistic (Cochran's Q) = 416.6828 on 136 degrees of freedom, (p-value = 0.0000). $I^2$ = 67.4%. F statistic = 44.0.
cML: constrained maximum likelihood; IVW: Inverse variance weighted; MIS: Melanoma in situ; MM: Invasive Melanoma; MR: Mendelian Randomization; RAPS: Robust adjusted profile score

## Discussion

The promise of early detection is that sooner removal of a tumor should result in overall decreased morbidity and mortality. Our challenge with invasive melanoma is that dysplastic nevi and melanoma in situ may never progress to clinically meaningful disease.^5^ Overdiagnosis or the increase in the diagnosis of such low-risk disease - a phenomenon more common in those with higher SES - may even result in more harm than good.^7^ Although much literature supports the hypothesis of melanoma overdiagnosis^2–6,8^, causal evidence for this phenomenon is outstanding. In this genetic correlation study, we provide three complementary lines of evidence to demonstrate causality between SES and MIS diagnosis. We identify a positive and significant overlap in the genetic architectures of SES and MIS but not SES and MM. We demonstrate that the genetic correlation between SES and MIS is consistent, even when focusing on the genetic architecture of MIS after subtracting that of MM. Finally, we apply MR and find further causal evidence for the effect of SES on MIS but not MM. All three of our complementary approaches examine different aspects of the genetic architectures underlying these traits and so the consistent results bolster confidence in our findings. Our results demonstrating causality between SES and MIS are consistent with an overdiagnosis of MIS, with a plausible mechanism being higher SES leading to more access to dermatological care and more diagnoses of MIS.

A particular strength of this study is the use of MM as a comparator condition. Any confounding or indirect contribution of SES’s genetic architecture to melanoma etiopathogenesis – for example, increased opportunities for outdoor leisure leading to more ultraviolet exposure^29^ or increased height resulting in more nevi^30^ – can be reasonably expected to equally impact MIS and MM. Thus, our findings support an extrinsic causal pathway, unless, of course, there exist biological differences between MIS and MM.^31^ This is an active area of investigation.

Our identification of a single variant, rs77270200, both highly significantly associated with increased melanoma risk and decreased SES is novel. rs77270200 is an intronic variant in GAS8, a gene involved in ciliary function.^32^ Interestingly, the association of this variant with income appears to be independent from its association with educational attainment.^10^ Hypothesizing on the mechanism by which this variant could influence melanoma is challenging given its high level of pleiotropism (Supplementary Table 1), but notable associations include pigmentary traits and non-melanoma skin cancers. Importantly, rs77270200’s association with both income and pigmentation related traits could potentially represent uncontrolled population stratification in the income GWAS.^10^ However, even if the biological interpretation of income’s genetic architecture is limited by population stratification, the fact that it statistically captures income differences allows for its use as a proxy for investigating causality between SES and MIS. In addition, our MR results supporting causality are robust to excluding this variant from the analyses (Supplementary Table 2).

Our results highlight that the genetic architecture underlying complex traits like disease phenotypes are not independent of the social context leading to their ascertainment. As a result, caution must be taken during the development and deployment of genetics-based clinical risk stratification tools like polygenic risk scores in order avoid exacerbating existing biases and disparities. Similarly, use of these genetic tools for research should account for, or at the very least acknowledge, this bias as a limitation. Whether the development of molecular diagnostic technologies can reduce this bias is a promising area for future research.

There are very important limitations of this study. The GWAS summary statistics we analyze are derived from individuals with primarily European genomic ancestries, thereby limiting generalization. While individuals with these ancestries typically have higher risk for melanoma^33^, investigations aimed at replicating our results using genetic data from diverse populations should be attempted as these data and analysis methods become available. Another limitation is that the genetic architecture of income explains only approximately 7% of the heritability of this trait and much of it reflects indirect rather than direct genetic effects.^10^ Despite this, the study reporting income’s genetic architecture identified several expected genetic correlations with health-related traits (Figure 4 from Kweon et al. 2025)^10^, bolstering confidence in our approach and findings.

## Conclusions

In this genetic association study, we identify that the genetic architecture of SES correlates with that of MIS but not MM and provide evidence for causality between SES and MIS. Our findings support overdiagnosis of MIS and demonstrate that disease genetic risk architecture is not inherently independent of social determinants of diagnosis. Thus, caution should be taken during the development and deployment of genetics-based clinical risk stratification tools.

## Supplementary Figures

**Supplementary Figure 1.**
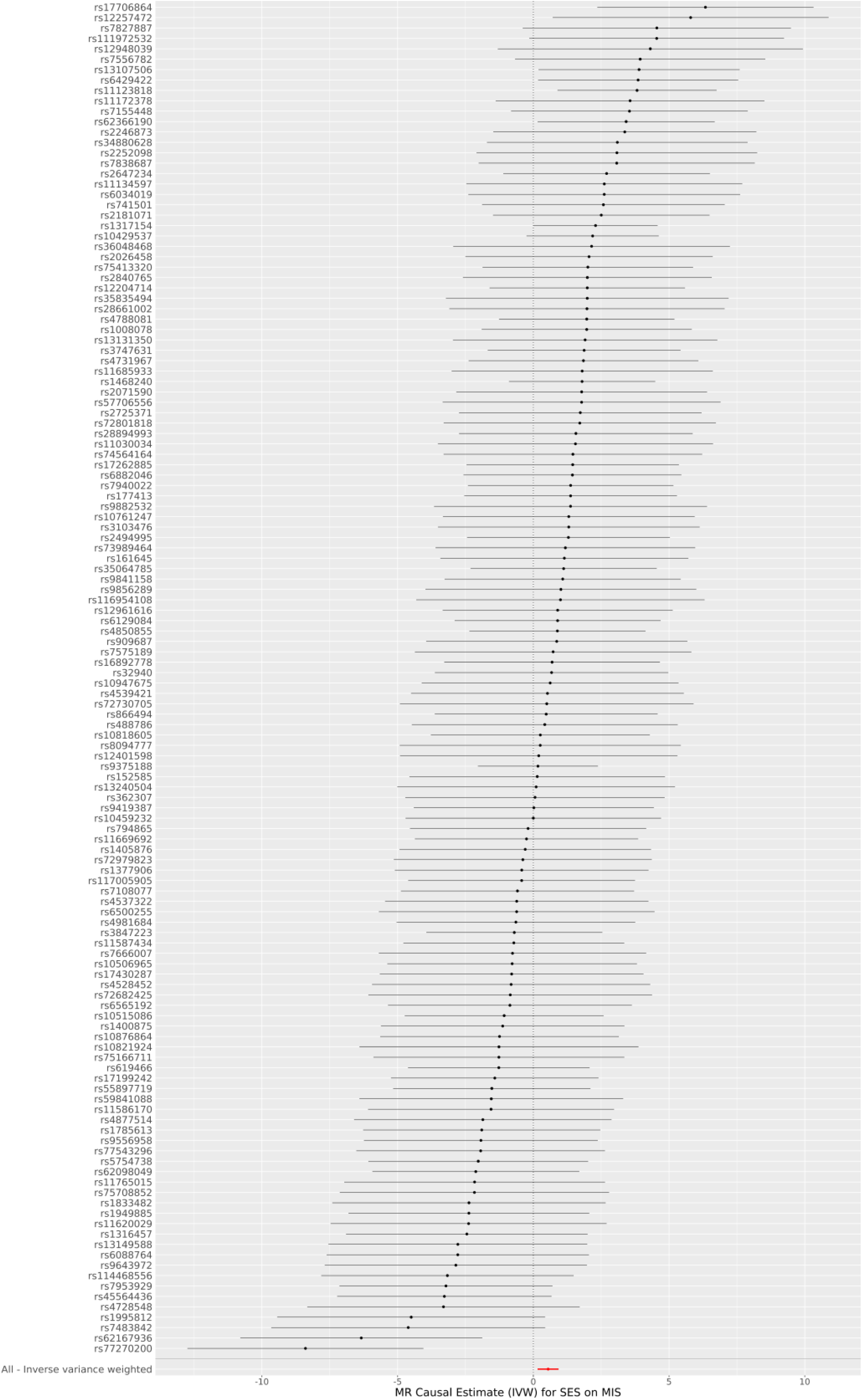
Single Genetic Variant Mendelian Randomization Inverse Variance Weighted Estimate for Socioeconomic Status on In situ Melanoma

**Supplementary Figure 2.**
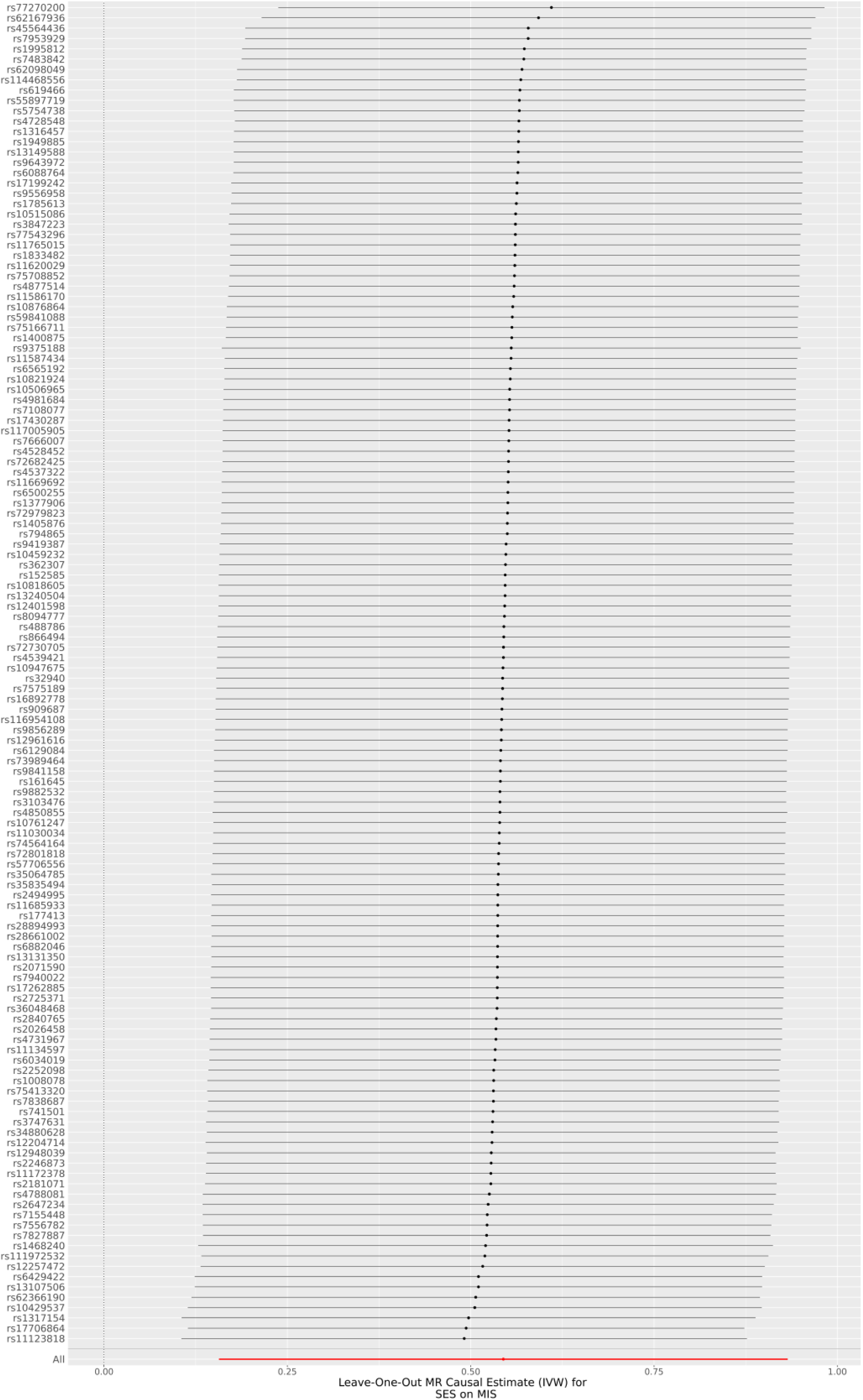
Leave-One-Out Mendelian Randomization Estimate for Inverse Variance Weighted Socioeconomic Status on In situ Melanoma

**Supplementary Figure 3.**
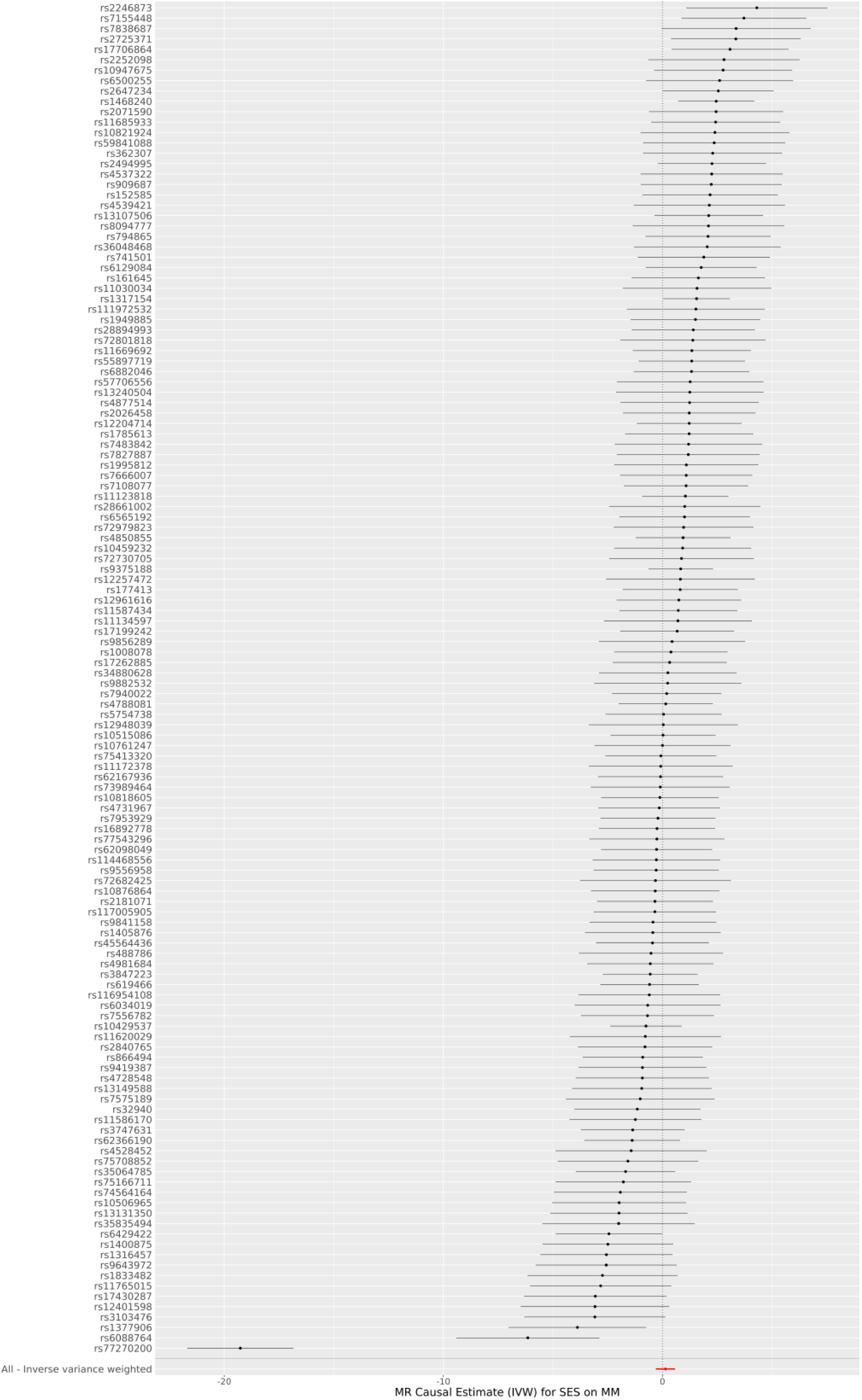
Single Genetic Variant Mendelian Randomization Inverse Variance Weighted Estimate for Socioeconomic Status on Invasive Melanoma

**Supplementary Figure 4.**
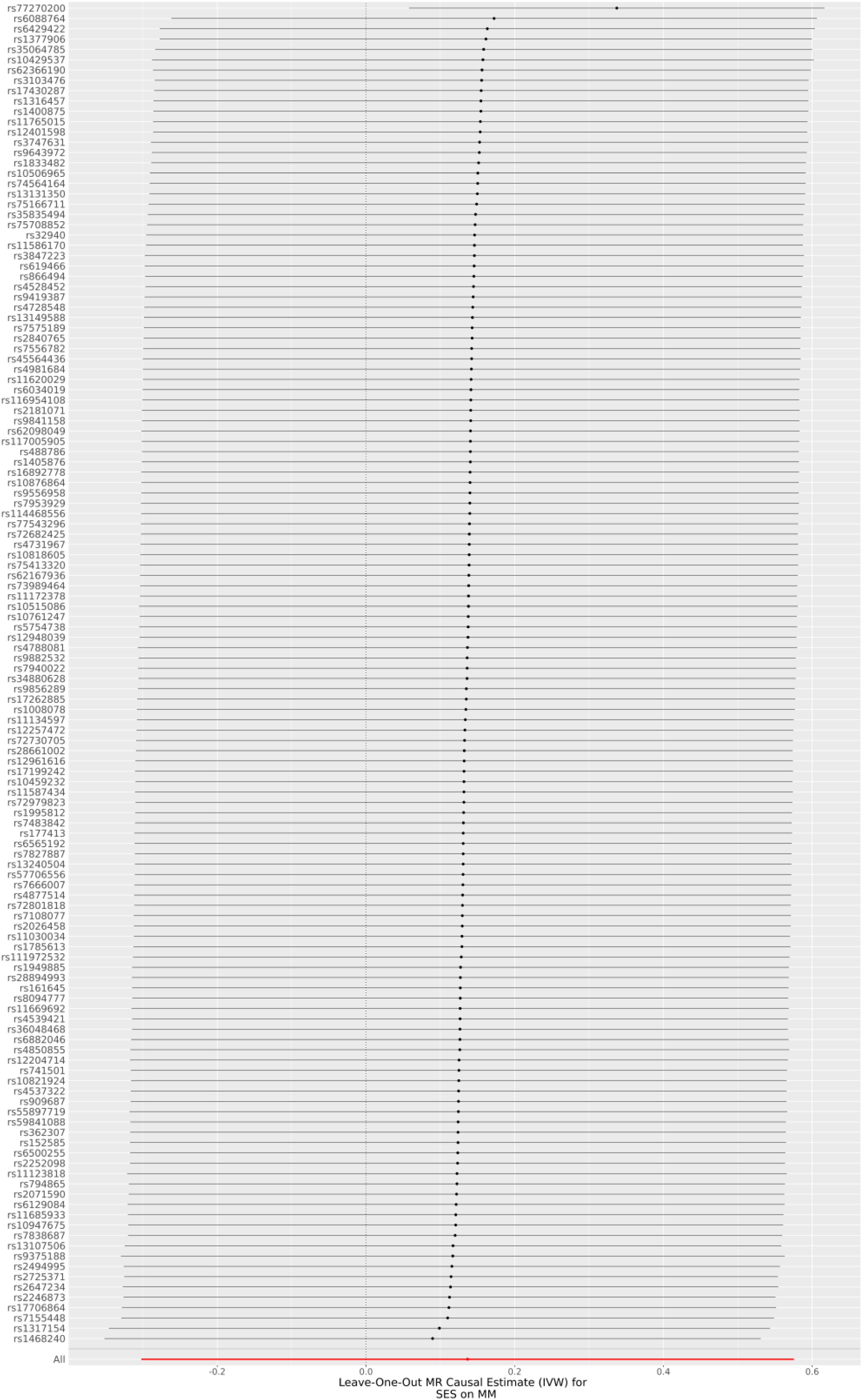
Leave-One-Out Mendelian Randomization Inverse Variance Weighted Estimate for Socioeconomic Status on Invasive Melanoma

## Data Availability

All data produced in the present study are available upon reasonable request to the authors

https://zenodo.org/records/12772644

https://nodes.desci.com/dpid/149

